# EPIDEMIOLOGICAL PROFILE OF CHLAMYDIA TRACHOMATIS INFECTION IN LOW-INCOME PREGNANT WOMEN IN KINSHASA

**DOI:** 10.64898/2026.03.21.26348957

**Authors:** Olivier Ndzouebeng, Georges Lelo Mvumbi, Bive Zono, Doudou Malekita Yobi, Pius Zakayi Kabutu, Tite Minga Mikobi

## Abstract

Chlamydia trachomatis (CT) is a sexually transmitted infection (STI). In 2020, an estimated 128.5 million new CT infections were reported among adults worldwide. The global prevalence was 4.0% in women and 2.5% in men. Young adults are the most frequently infected. This bacterial STI is often asymptomatic but can lead to serious complications, particularly in pregnant women.

The objective of this study was to determine the prevalence and sociodemographic profile of CT infection in pregnant women living in resource-limited settings.

**Methods:** We conducted a cross-sectional study between June 2023 and december 2023 in a maternity ward located west of Kinshasa in an impoverished area with a low-income population. During the study period, we collected 239 cervical swab samples from pregnant women. CT DNA was extracted using the QIAamp DNA Mini Kit. Molecular diagnosis was performed by amplification of a 201 bp fragment of bacterial 16S rDNA from the cryptic plasmid.

**Results:** The age group most affected by CT in our cohort was 25 to 35 years (65.83%); married women were more represented than single women (83.22% versus 16.77%). The incidence of CT infection was 18%. The most common vaginal symptoms associated with the infection were vaginal itching and abnormal vaginal discharge, while the most common hypogastric symptom was chronic pelvic pain. Prematurity and spontaneous abortions were the most frequently observed pregnancy complications.

**Conclusion:** Chlamydia trachomatis (CT) infection is common among pregnant women living in poverty in Kinshasa. The infection particularly affects the most sexually active age group. It is associated with vaginal and hypogastric symptoms, as well as complications related to the progression of the pregnancy.

## Introduction

Chlamydia are obligate intracellular bacteria. Three species are pathogenic to humans. Chlamydia pneumoniae is a cause of community-acquired pneumonia. Chlamydia psittaci is the cause of psittacosis, a zoonotic disease transmitted by birds. Chlamydia trachomatis causes trachoma and sexually transmitted and ocular infections. Chlamydia trachomatis (CT) infection can be transmitted during vaginal, anal, or oral sex [1]. This infection is often asymptomatic; however, it can be associated with non specific symptoms such as urethral and vaginal discharge and painful urination [1]. The risk of CT infection increases with the number of sexual partners. In 2020, the World Health Organization (WHO) estimated 128.5 million new CT infections worldwide [2]. This is a public health problem [3]. The most affected age group is between 15 and 49 years old [2]. This is also the most sexually active age group. The infection affects both men and women. In women of reproductive age, untreated CT infection is the leading cause of infertility. It is also responsible for chronic pelvic inflammatory disease (PID), salpingitis, and, in the long term, fallopian tube obstruction [4]. In pregnant women, CT infection is responsible for miscarriages, ectopic pregnancies, premature rupture of membranes, premature births, low birth weight, and sometimes stillbirth [5-7]. A pregnant woman can transmit the infection to her newborn. Infants born to mothers with Chlamydia trachomatis may develop an eye infection or pneumonia [1]. In the Democratic Republic of Congo (DRC), as in many sub-Saharan African countries, it is estimated that between 0 and 31% of pregnant women are infected with Chlamydia trachomatis [8-11]. This infection is the leading cause of infertility in young women, accounting for 95% of pregnancies in sub-Saharan Africa. Chlamydia diagnosis is primarily clinical; microbiological diagnosis can be performed using molecular techniques, but this requires significant infrastructure, which is rarely available in endemic areas.

The objective of this study was to determine the sociodemographic profile of chlamydia infection in pregnant women living in resource-limited settings.

## METHODS

We conducted a cross-sectional study between June 2023 and december 2023. The study was carried out in a maternity ward located in the western part of Kinshasa. Due to its geographic location, the maternity ward is situated in a neighborhood where the majority of households live with limited resources. The delivery rate at this maternity ward is the highest in Kinshasa, averaging 120 deliveries per month. In this part of Kinshasa, sexual promiscuity is very high among unmarried individuals. The level of education is low, and the population’s income is among the lowest in Kinshasa. During the study period, we collected 239 endocervical swab samples from the cervix of pregnant women.

### Inclusion criteria

In this study, we included all pregnant women who consulted a doctor during the study period and who signed the informed consent form.

### Non inclusion criteria

In this study, we did not include pregnant women who received antibiotic treatment two weeks prior to sample collection. We also did not include pregnant women who had sexual intercourse within 48 hours of sample collection.

### Exclusion criteria

We excluded from the study all samples with a bacterial DNA concentration below 95 µg/µl and those with a nanodrop absorbance below 260/280, corresponding to an average purity of 1.94.

### Sample Collection

Swab samples were collected during prenatal consultations. With the pregnant woman in the gynecological position, we used standard equipment for a gynecological examination. The sterile swabs for collection and the transport medium were supplied by COPAN eNAT® FLOQSwabs® MiniTip 551C.

### Sampling Technique

With the pregnant woman in the gynecological position, we performed vulvoperineal disinfection. Then, we inserted the speculum and disinfected the vagina and cervix with a mild disinfectant. Finally, we inserted the sterile swab into the endocervix. We gently rotated the swab for 10 seconds within the endocervical canal of the uterus. We then withdrew the swab, avoiding contact with the vaginal mucosa. The swab was inserted into the tube containing the aforementioned transport medium. Each sample was identified before transfer to the laboratory.

### Laboratory DNA Extraction

In the laboratory, we extracted bacterial 16S rDNA from the cryptic Chlamydia trachomatis plasmid. The DNA was extracted using a QIAamp DNA Mini Kit. The extraction procedure was as described by the manufacturer in the Artus C. trachomatis Plus RG PCR Kit manual. After bacterial DNA extraction, DNA quality control was performed using the NanoDrop. All samples with a concentration below 95 ng/µl and a mean purity below 1.94 were excluded from the study.

### Chlamydia trachomatis PCR

Chlamydia DNA amplification was performed using an ABI Veriti 96 Well thermocycler. Our primers were provided by Eurofins Scientific: CTP1 (5’-TAGTAACTGCCACTTCATCA-3’) and CTP2 (5’-TTCCCCTTGTAATTCGTTGC-3’). These primers allowed us to amplify the 101 bp fragment of interest located on the bacterial 16S rDNA of the cryptic Chlamydia trachomatis plasmid. Thermo Scientific™ DreamTaq Green PCR Master Mix (2X) was used according to the manufacturer’s recommendations.

### Amplification Program

Our amplification program consists of three traditional PCR steps: initiation or denaturation at 95°C for 5 minutes, hybridization at 55°C for 1 minute, and elongation at 72°C for 1 minute. The total number of cycles was 35.

### Post-PCR and Positive Diagnosis of Chlamydia trachomatis

The PCR product (amplicons) was migrated on a 2% agarose gel. Each amplicon run consisted of one molecular weight marker well, one positive control sample, one negative control sample (PCR water), and the analysis samples. The positive control used in this study was provided by Eurofins Scientific. The sample was declared positive when, after amplification, the amplification product showed the 101 bp band on the agarose gel at the same level as the positive control. This band confirms the presence of the bacterial 16S rDNA of the cryptic Chlamydia trachomatis plasmid. Pregnant women with a positive CT diagnosis were informed and managed by the national sexually transmitted infection control program. Figure-1 below shows an example of a positive sample from one of our analysis runs.

**Fig-1:**
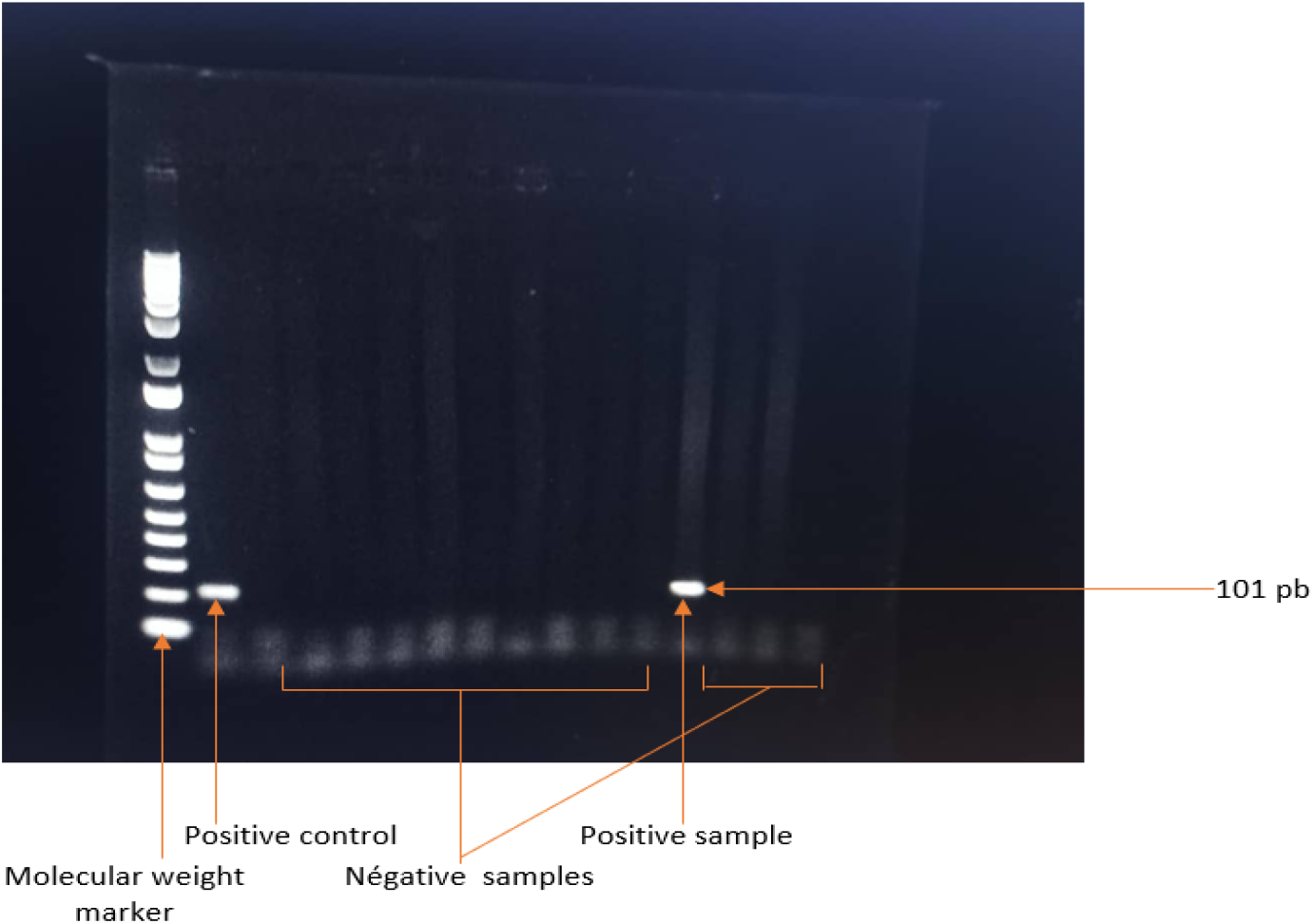
positive PCR test for chlamydia trachomatis

### Statistics

The data for this study were entered into Excel and then exported to EPIINFO version 7. The distribution was normal; we calculated the means and standard deviations. The prevalence of C. trachomatis was expressed with a 95% confidence interval. Risk factors were assessed using bivariate and univariate logistic regression. A p-value < 0.5 was considered statistically significant.

## Results

### Final Study Size Design

After DNA concentration analysis using nanodrops, we selected 161 pure samples with an absorbance of 260/280 and an optimal concentration of 95 ng/µl.

Table 1 above presents the distribution of pregnant women according to the sociodemographic characteristics studied. This table shows that the most represented age group in our series was 25 to 35 years (65.83%); married women were more represented than single women (83.22% versus 16.77%); the trade profession was the most represented at 40.99%; finally, pregnant women with a secondary education level were the most represented (83.2%).

**Table 1:** Distribution of pregnant women according to sociodemographic characteristics.

| Sociocultural variables |  | Total pregnant women ( N = 161) | Frequencies (%) |
| --- | --- | --- | --- |
| Age (years) | < 25 years | 29 | 18,01% |
|  | 25 to 35 years | 106 | 65,83% |
|  | >35 years | 26 | 16,14% |
| Marital status | Single | 27 | 16,77% |
|  | Bride | 134 | 83,22 |
| Occupation | Household | 39 | 24,22% |
|  | Student | 4 | 2,48% |
|  | official | 12 | 7,45% |
|  | Trader | 66 | 40,99% |
|  | Unemployed | 30 | 18,63% |
|  | Other | 10 | 6,21% |
| Level of studies | Illiterate | 1 | 0,5% |
|  | Primary | 3 | 1,86% |
|  | Secondary | 134 | 83,22% |
|  | University | 23 | 14,28% |

Table 2 shows the distribution of pregnant women according to their gynecological and obstetric status. We note that multiparous pregnant women were more represented (49.06%), 12.42% of pregnant women had a history of spontaneous abortion, 3.72% of pregnant women had a history of a sexually transmitted infection, and 7.45% had experienced a threatened premature birth.

**Table 2:** Distribution of pregnant women according to parity and gynecological status.

| Gyneco-obstetric characteristics | Total pregnant womenI (N = 161) | Frequencies (%) |
| --- | --- | --- |
| Nulliparous | 33 | 20,49% |
| Primiparous | 49 | 30,43% |
| Multiparous | 79 | 49,06% |
| spontaneous abortions | 20 | 12,42% |
| history of sexually transmitted infection | 6 | 3,72% |
| history of threatened miscarriage | 12 | 7,45% |

Table 3 above shows the distribution of positive chlamydia PCR tests in the study population according to the sociocultural characteristics of the pregnant women. We observed that 18% of the pregnant women in our study cohort were infected with Chlamydia trachomatis. The most infected age group was 25 to 35 years (55.9%), with married women being more infected than single women. Finally, pregnant women with a secondary education were more infected (14.9%) than the others.

**Table 3:** Distribution of positive chlamydia PCR in the study population according to the sociocultural characteristics of pregnant women.

| Sociocultural characteristics of pregnant women | Chlamydia PCR positive (n = 29) (18%) | Chlamydia PCR negative (n = 132) (82%) | p |
| --- | --- | --- | --- |
| Age (years) |  |  | 0,34 |
| <25 years | 6 (3,72%) | 23 (14,28%) |  |
| 25 to 35 years | 16 (8,93) | 90 (55,9%) |  |
| >35 years | 7 (4,34%) | 19 (11,8%) |  |
| Marital status |  |  | 0,56 |
| Single | 24 (14,9%) | 110 (68,32%) |  |
| Bride | 5 (3,1%) | 22 (13,66%) |  |
| Level of studies |  |  | 0,77 |
| Illiterate | 0 (0%) | 1 (0,6%) |  |
| Primary |  |  |  |
| Secondary |  |  |  |
| University |  |  |  |

Figure 3 above shows the frequency of symptoms and complications associated with Chlamydia trachomatis infection in the study population. It highlights the symptoms commonly associated with CT, and certain pregnancy complications were observed more frequently in pregnant women with a positive CT PCR test.

## DISCUSSION

Chlamydia trachomatis causes trachoma and sexually transmitted and ocular infections. Untreated chlamydia trachomatis infection is the leading cause of infertility in women and is responsible for chronic pelvic inflammatory disease. This infection constitutes a public health problem. The objective of our study was to determine the prevalence, epidemiological profile, and clinical characteristics of chlamydia trachomatis infection in pregnant women living in resource-limited settings.

Our study showed that 18% of pregnant women in our study population were infected with Chlamydia trachomatis, and the most affected age group was 25 to 35 years. Our prevalence is lower than that reported by Manca et al. [12], who found a prevalence of 24% in a resource-limited setting like ours. This difference may be attributed to the fact that in the Manca et al. study, they investigated the prevalence of two sexually transmitted infections, TC and Neisseria gonorrhea. However, our prevalence of 18% is higher than the 2% prevalence reported by Torrone in the USA [13, 14]. This difference could be attributed to the standard of living of the Torrone study population. Several studies report low prevalences between 3% and 9% [6, 17, 18] compared to our study. We believe this difference could be due to the diagnostic technique used. Indeed, in these studies with low prevalence, the diagnostic technique used was cell culture; whereas in our study, we used molecular testing. The most affected age group in our study was 25 to 35 years old. Our observation is consistent with many studies that report this age group as being most affected by CT infection because it represents the sexually active middle age. In our study, married women were the most affected category by chlamydia trachomatis infection. This observation is due to the fact that our target population consisted of pregnant and sexually active women rather than single women. The secondary education level was the most affected in our series. This observation is also due to the fact that in sub-Saharan Africa, early marriage prevents girls from pursuing university studies. Symptomatically, the clinical symptoms associated with CT infection and certain pregnancy complications, such as prematurity and miscarriages, were observed more frequently in pregnant women infected with CT. This observation confirms data from the literature indicating that CT infection is associated with the vaginal and pelvic symptoms and pregnancy complications described in Figure 2 of our results.

**Fig-2:**
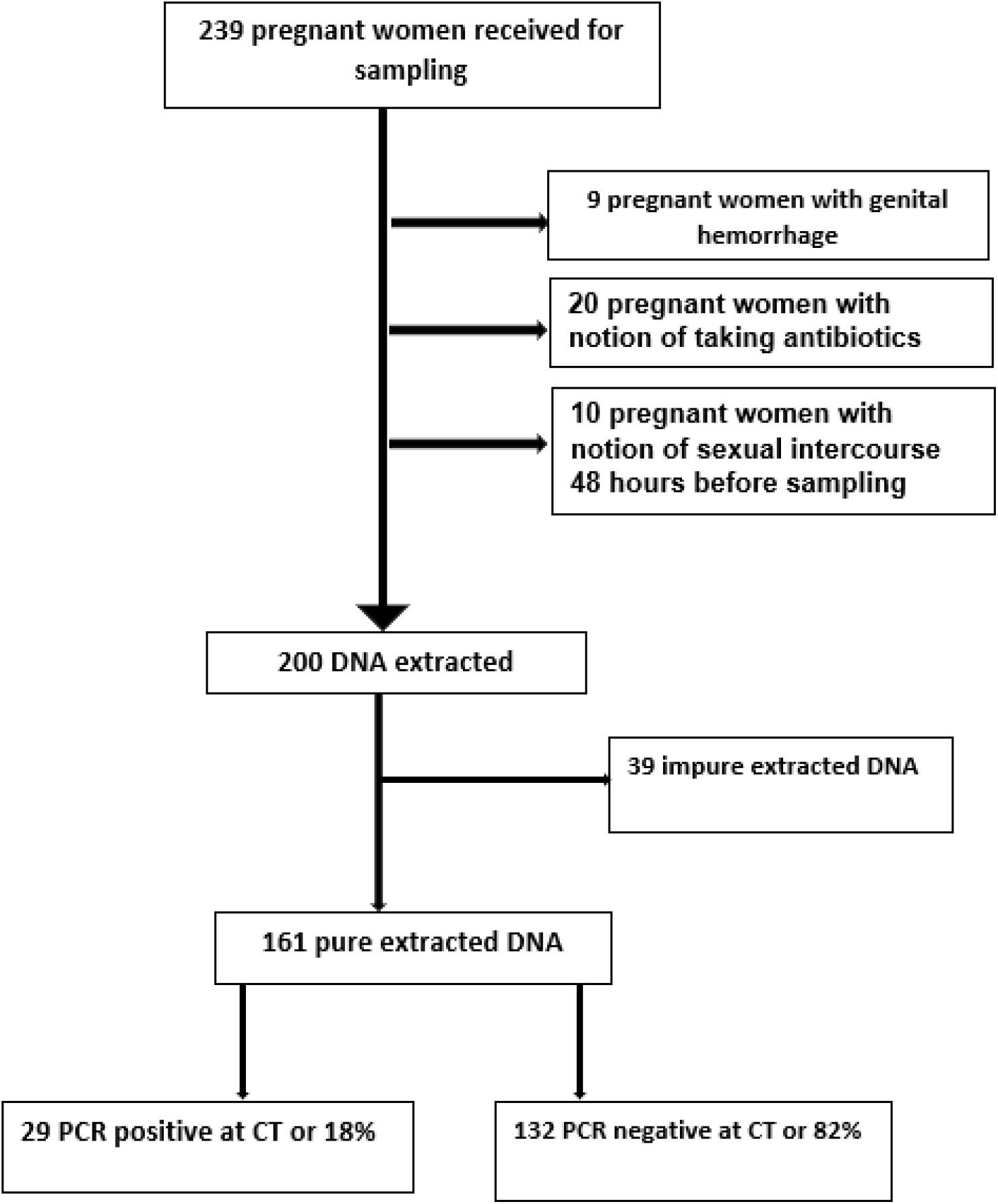
Study Population Design

**Fig-3:**
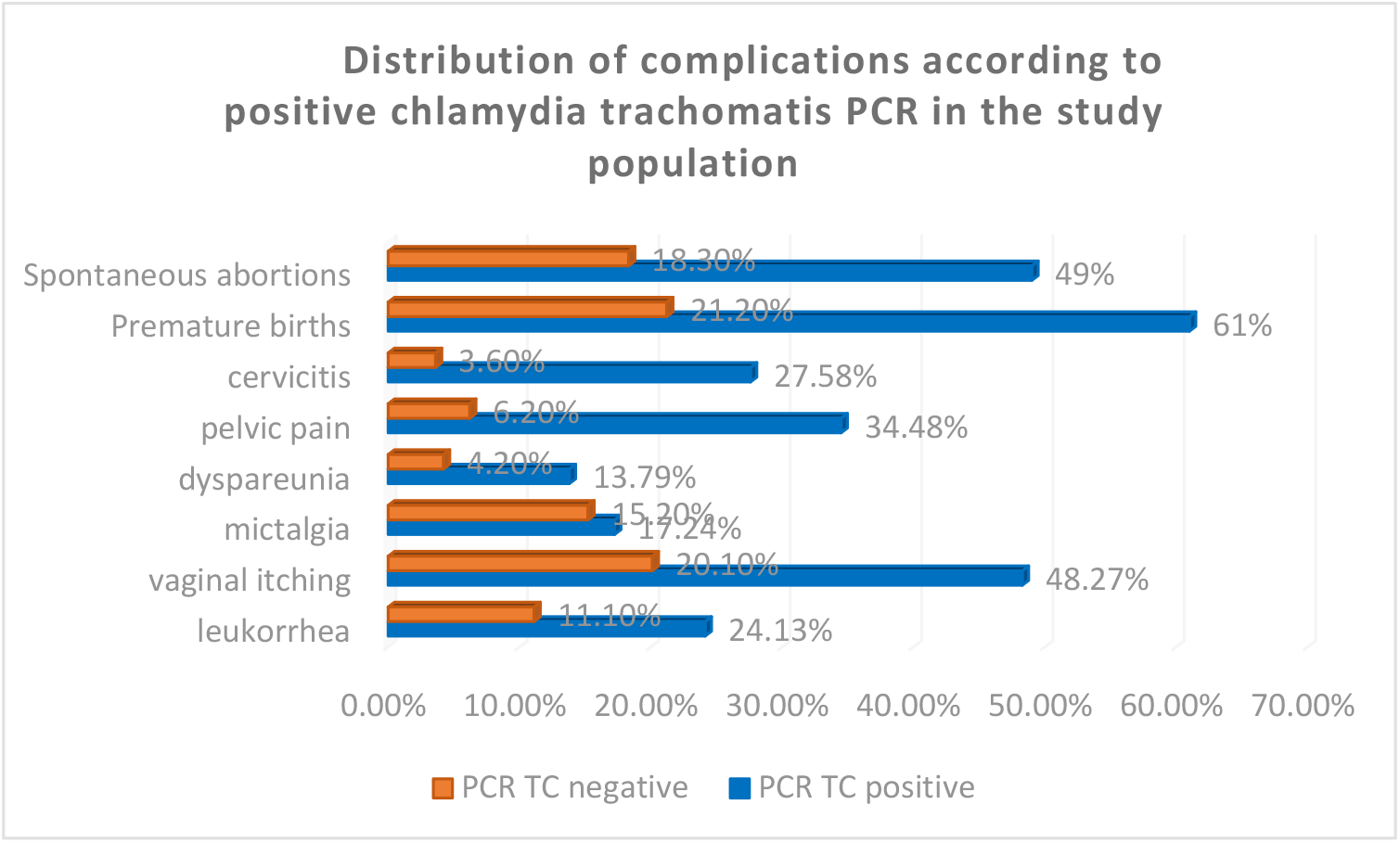
Frequency of complications and symptoms in pregnant women with a positive Chlamydia trachomatis PCR test

## Conclusion

Chlamydia trachomatis infection in pregnant women in resource-limited settings is a public health challenge. The infection often progresses asymptomatically, making early diagnosis difficult. Early screening should be recommended for at-risk individuals. In the absence of systematic screening, the infection persists and progresses undetected, exacerbated by limited access to rapid diagnostic tests in sub-Saharan Africa.

## Acknowledgements

We sincerely thank the participants in this study, and we also thank the national program for the fight against sexually transmitted infections for the free care provided to pregnant women who were screened.

## Funding sources

this study has not received funding from any institution, government or donor

## Conflict of interest

the authors declare no conflict of interest

## Data availability

All data relating to this manuscript are available upon request from the authors.

## Ethical approval

The study was approved by the ethical committee of the school of public health of the University of Kinshasa (Approval reference: ESP/CE/08/2023), DRC. Informed consent was obtained from all patients before their inclusion in the study.

## Authors’ contribution

**ON**: data collection, sample analysis, interpretation of results; **GLM**: interpretation of results, correction of the manuscript and supervision of data collection; **BZ** : development of the PCR program, PCR analysis; **DMY**: development of the PCR program, PCR analysis; **PZK**: development of the PCR program, PCR analysis; **TMM:** topic design, manuscript writing, data collection.

## What does this study add?

This study is the first RDC to determine the prevalence of CT using molecular diagnostics.

## Notes

### Competing Interest Statement

The authors have declared no competing interest.

### Funding Statement

The author(s) received no specific funding for this work.

